# China’s effective control and other countries’ uncharted challenge against COVID-19: an epidemiological and modelling study

**DOI:** 10.1101/2020.04.28.20083899

**Authors:** Lingling Zheng, Qin Kang, Weiyao Liao, Xiujuan Chen, Shuai Huang, Dong Liu, Huimin Xia, Jinling Tang, Huiying Liang

**Affiliations:** Department of Clinical Data Center, The Guangzhou Women and Children’s Medical Center, Guangzhou Medical University, Guangzhou, Guangdong, China; Department of Nutrition, School of Public Health, Sun Yat-sen University, Guangzhou, China; The Guangdong Provincial Children’s Medical Research Center, Guangzhou, Guangdong, China; Department of The School of Public Health and Primary Care, Chinese University of Hong Kong, Hong Kong, China

## Abstract

**Background:** On the present trajectory, COVID is inevitably becoming a global epidemic, leading to concerns regarding the pandemic potential in China and other countries.

**Objective:** In this study, we use the time-dependent reproduction number (R_t_) to comprise the COVID transmissibility across different countries.

**Methods:** We used data from Jan 20, 2019, to Feb 29, 2020, on the number of newly confirmed cases, obtained from the reports published by the CDC, to infer the incidence of infectious over time. A two-step procedure was used to estimate the R_t_. The first step used data on known index-secondary cases pairs, from publicly available case reports, to estimate the serial interval distribution. The second step estimated the R_t_ jointly from the incidence data and the information data in the first step. R_t_ was then used to simulate the epidemics across all major cities in China and typical countries worldwide.

**Results:** Based on a total of 126 index-secondary cases pairs from 4 international regions, we estimated that the serial interval for SARS-2-CoV was 4.18 (IQR 1.92 – 6.65) days. Domestically, R_t_ of China, Hubei province, Wuhan had fallen below 1.0 on 9 Feb, 10 Feb and 13 Feb (R_t_ were 0.99±0.02, 0.99±0.02 and 0.96±0.02), respectively. Internationally, as of 26 Feb, statistically significant periods of COVID spread (R_t_ >1) were identified for most regions, except for Singapore (R_t_ was 0.92±0.17).

**Conclusions:** The epidemic in China has been well controlled, but the worldwide pandemic has not been well controlled. Worldwide preparedness and vulnerability against COVID-19 should be regarded with more care.

What is already known on this subject?
- The basic reproduction number (R0) and the-time-dependent reproduction number (Rt) are two important indicators of infectious disease transmission. In addition, Rt as a derivative of R0 could be used to assess the epidemiological development of the disease and effectiveness of control measures. Most current researches used data from earlier periods in Wuhan and refer to the epidemiological features of SARS, which are possibly biased. Meanwhile, there are fewer studies discussed the Rt of COVID-19. Current clinical and epidemiological data are insufficient to help us understand the full view of the potential transmission of this disease.

What this study adds?
- We use up-to-data observation of the serial interval and cases arising from local transmission to calculate the Rt in different outbreak level area and every province in China as well as five-top sever outbreak countries and other overseas. By comparing the Rt, we discussed the situation of outbreak around the world.

## Introduction

### Background

The latest threat to global health is the ongoing outbreak of SARS CoV-2 (formerly 2019-nCoV) infection emerged in December 2019 in Wuhan, Hubei Province, China.^1^ As of 4 March, 2020, there had been 80,422 confirmed cases including 2,984 deaths in China and over 12,668 confirmed cases in other 76 countries.^2^ Local transmission outside China has been reported in both developed countries^3^ and developing countries.^4^

The Chinese government has instituted restrictions on population movements with an eye toward slowing the spread of this new disease, such as Wuhan and nearby cities in Hubei province lockdown approaches, large-scale quarantine,^5^ public entertainment places closure, banning public gathering,^6^ and mandatory wearing of masks in public places.^7^ Subsequently, the number of new cases reported each day has plummeted the past few weeks. However, the opposite has happened in other countries (Fig 1). Just this past week, the number of affected countries shot up from 37 to 61 and the number of new cases reported each day are growing exponentially. Thus, whether China’s current control efforts are enough or whether worldwide additional interventions are required remains an open question.

**Figure 1.**
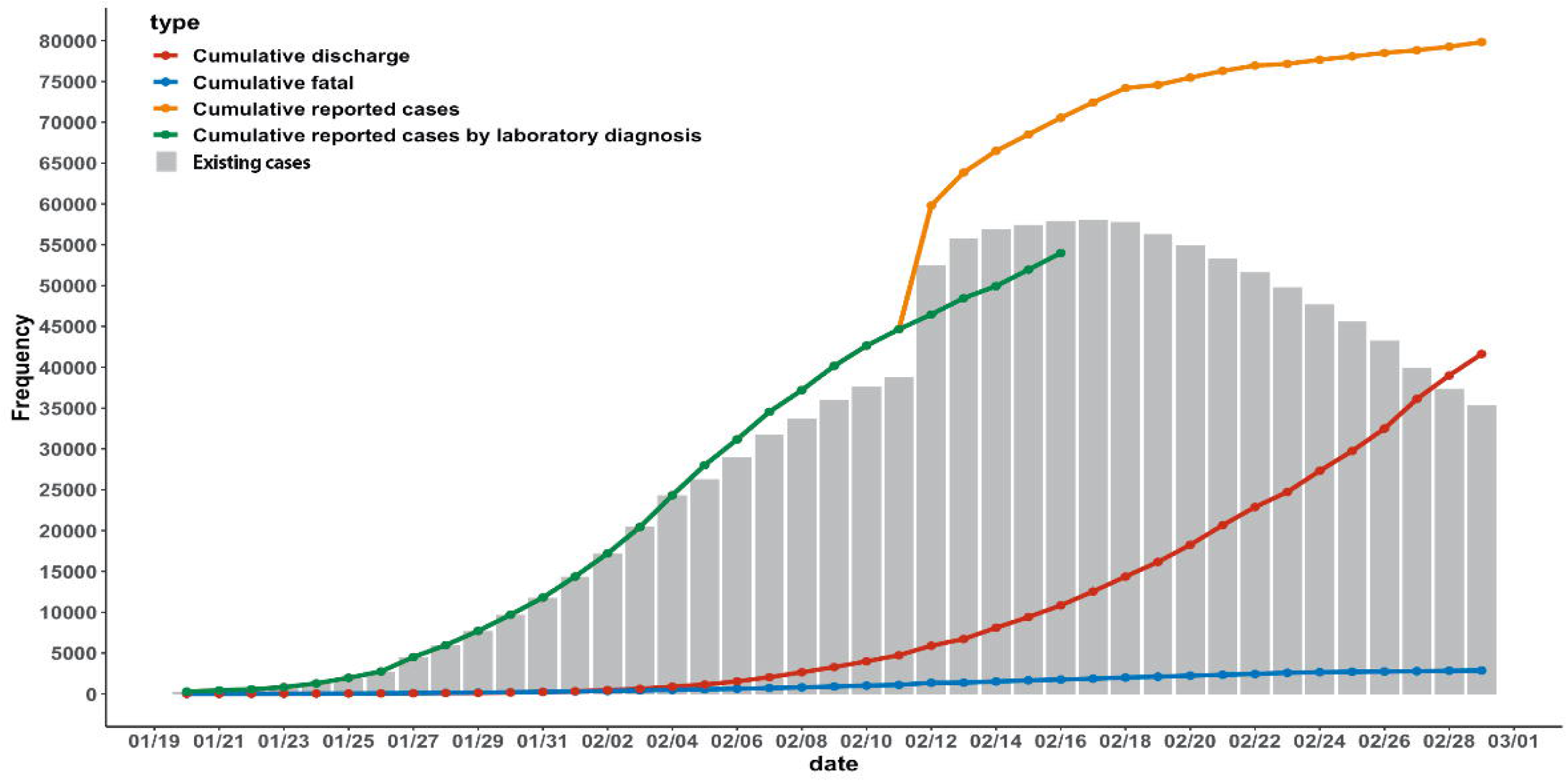
The distribution of cumulative report cases, fatal case, discharge and exiting cases in China. Four lines showed the distribution of cumulative reported cases, cumulative reported cases by laboratory diagnosis, fatal and discharge in China. And the bar chart showed the daily exiting cases in hospital in China.

There is a proxy, however, that could help to evaluate disease progression and that have not been sufficiently explored in measuring COVID-19 transmissibility across different countries. The time-dependent reproduction number (R_t_)^8^ represents the expected number of secondary cases arising from a primary case infected at time *t*, which changes throughout the outbreak. The R_t_ is directly related to the type and intensity of interventions to control an epidemic. If the value of R_t_ is and remains >1, a sustained outbreak is likely. While R_t_ is <1, the outbreak will die out. The aim of control interventions is typically to reduce the reproduction number below one as soon as possible.

Particularly, Thompson developed a novel framework to estimate the R_t_ based on the approach developed by Wallinga. Rather than depending on the previous estimates of the serial interval, the distribution of which may be unavailable early in an outbreak or with significant uncertainty, this new framework allowed the direct estimation of the serial interval through integrating information from known pairs of index and secondary cases. They applied this method to data from H1N1 influenza in the USA in 2009 and 2013 Ebola virus epidemic, showing improvements of the precision and accuracy of R_t_.

### Objectives

In this study, we used the statistical framework of Thompson *et al.^8^* to describe and estimate the domestic and international ongoing control situation of the epidemic based on officially reported COVID case data.

## Methods

### Data Collection

Firstly, we collected the daily cumulative confirmed, fatal and discharge cases data from Chinese CDC’s official website as of February 29, 2020.^9^ Then, we collected 126 index-secondary infector-infected pairs of confirmed cases with known or coarsely reported date of symptom onset by scanning publicly available case reports from official health websites or reliable news or other published papers as of February 16, 2020 (S1 Table).^4,10^ Last, we further collected the daily new confirmed cases overseas from WHO reports as of February 26, 2020.^2^

### Process of Data Analysis

#### Epidemic curve

The epidemic curve was constructed by date of cumulative reported cases by laboratory diagnosis, cumulative reported all cases (including laboratory diagnosis and clinical diagnosis), cumulative fatal, cumulative discharge and existing cases, and key dates relating to epidemic identification.

#### The serial interval

The serial interval was the period of time between analogous phases of an infectious illness in successive cases of a chain of infection that is spread from person to person.^11^ We performed Bayesian parametric estimation of the serial interval distribution from 126 pair data using data augmentation Markov chain Monte Carlo (MCMC). And we fitted a gamma distributed serial interval distribution offset by one day.^12^ The MCMC estimation was used to a joint posterior sample data for the parameters of the gamma distribution, with the parameter set corresponding to a step in the MCMC chain.

#### The time-dependent reproduction number (R_t_)

The time-dependent reproduction number, R_t_, was an important parameter for assessing whether current control efforts were effective or whether additional interventions were required.^13^ The value of R_t_ represented the expected number of secondary cases arising from a primary case infected at time t. This value changed throughout an outbreak. If the value of R_t_ was below one, the outbreak would die out. However, while R_t_ was larger than one, a sustained outbreak was likely. The aim of control interventions is typically to reduce the reproduction number below one.^11^ We used R.N. Thompsona’s method^8^ to estimate disease incidence time series and serial interval data for integrating the estimation of the serial interval distribution within the estimation of R_t_. This allows R_t_ to be estimated directly not only from the most up-to-date incidence data, but also from up-to-date serial interval data.

### Statistical Analysis

Analyses aimed to describe the change epidemic of COVID-19 in China and outside China. First, we described the overall epidemic curve in China. Second, 126 infector-infected pairs of confirmed cases characteristics were described, including age, gender and the relationship between infectors and the infected. We calculated the serial interval by 126 infector-infected pairs. Third, we assessed the geographical distribution of daily new confirmed cases by laboratory diagnosis across all 31 provinces in China, grouping into three regions with different outbreak levels: Wuhan city (the main epidemic area), Hubei province excluding Wuhan and other provinces in China excluding Hubei Province. And we compared the R_t_ among these three regions. Last, we used the same method to estimate the development of epidemics outside China, separating six groups: Japan, Singapore, South Korea, Italy, Iran (which were the five top most serious epidemic countries) and other outside countries. Data analyses were performed by the use of R software (version 3.6.0), with “EpiEstim” for estimating the serial interval and Rt.

## Results

### Overall epidemic trends of COVID-19

As of 29 February, 79,824 confirmed cases of COVID-19 in 32 provinces has been reported to the China CDC surveillance system, of which 2,870(3.60%) were fatal, 7,356(20.85%) were severe and 41,625(52.15%) were cured. The overall epidemic trends of NCIP were shown in Figure 1. On February 12, the cumulative reported cases split into two curves. That’s because on this day new clinically confirmed cases of COVID-19 were added in Hubei Province including Wuhan. Although the existing confirmed cases have been on the rise, they had gradually flattened in recent days. In addition, the number of discharged patients also showed a gradually increasing trend, the number of fatal remained increased but slowly. Therefore, the number of existing will begin to show a downward trend soon.

### Estimated the serial time

The characteristics of 126 infector-infected pairs of confirmed cases were showed in Table 1. We found out the relationship of 83 infector-infected pairs. A half of pairs were belonged to elder and junior. And nearly a third of pairs were couples. We fitted a gamma distributed serial interval offset by one day which was shown in Figure 2. The median serial time was 4.18 with IQR 1.92 to 6.65 days.

**Table 1.**
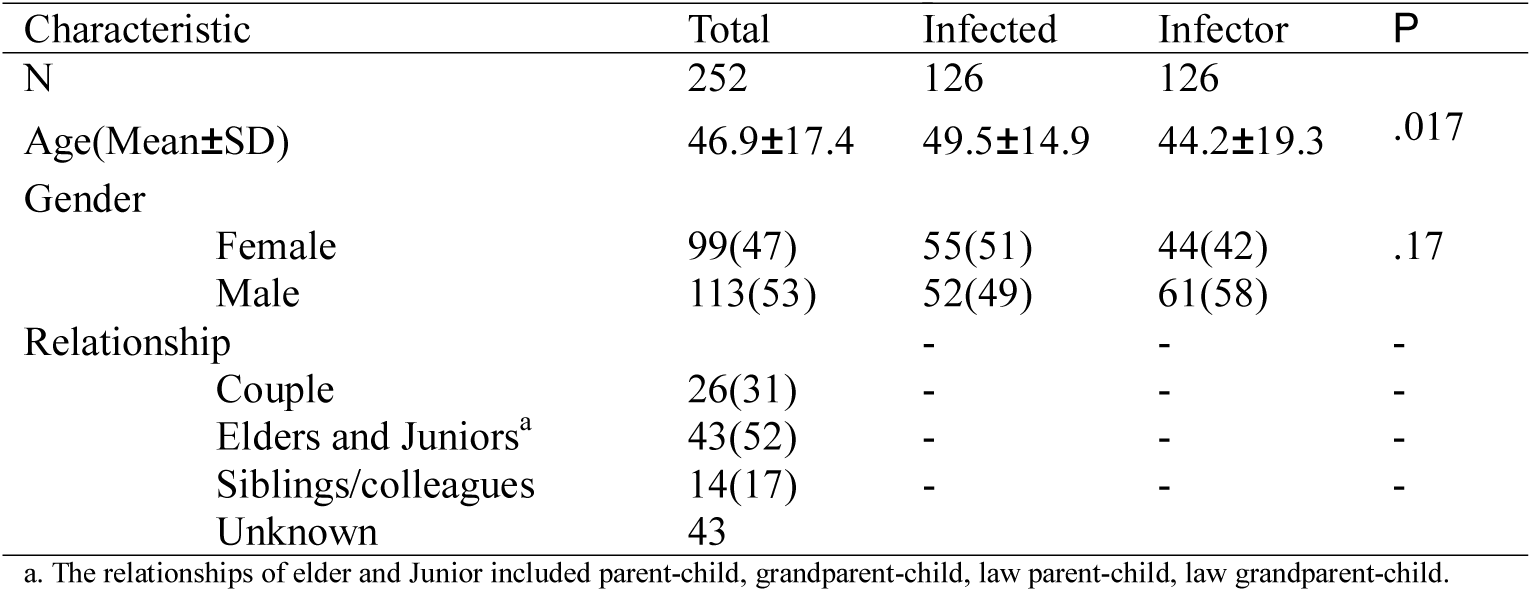
The characteristics of 126 infector-infected pairs of confirmed cases.

**Figure 2.**
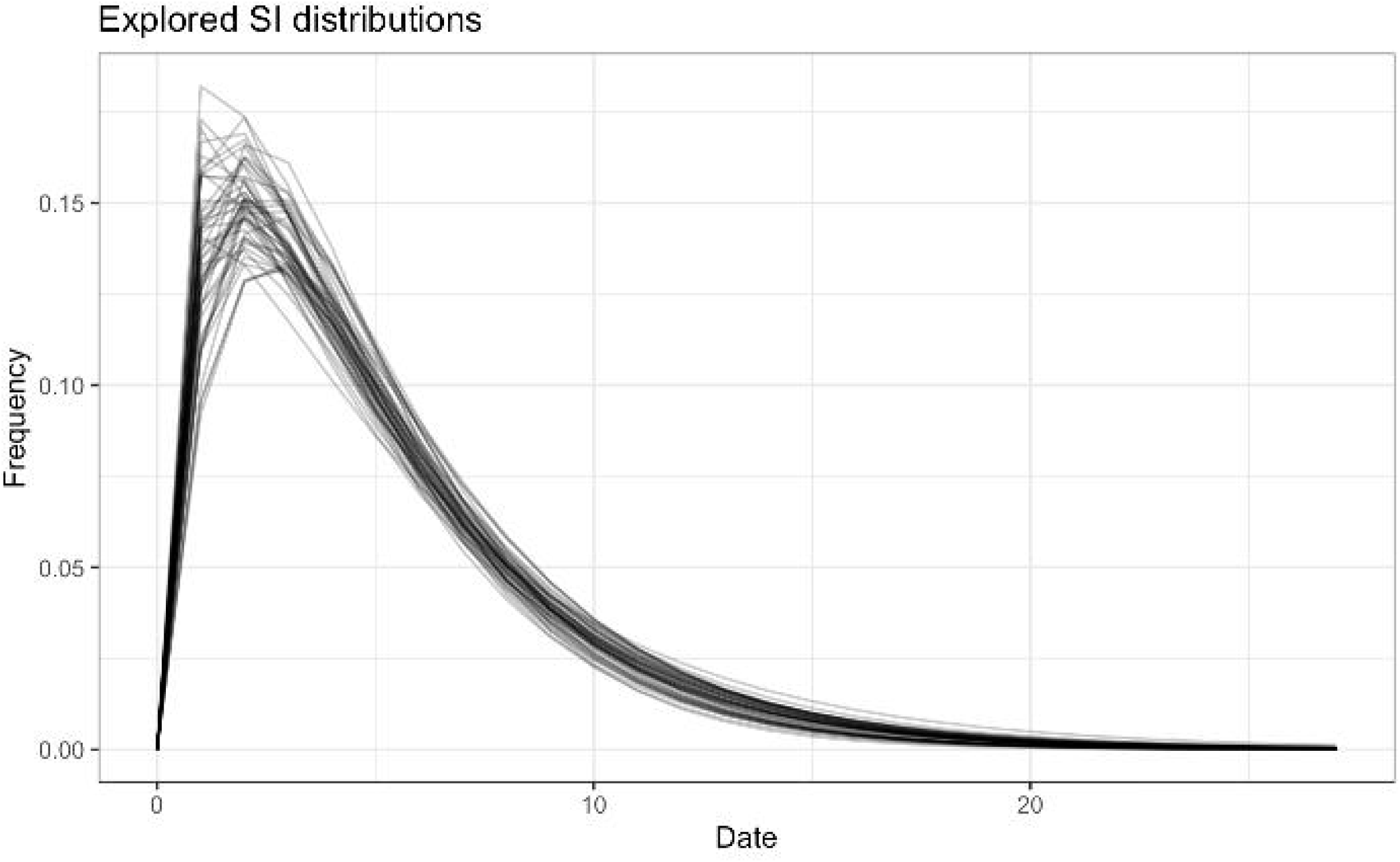
The distribution of serial interval time.

### Estimated R_t_ in three areas with varying levels of outbreak

Figure 3A. compared the daily incidence in three regions with different levels of outbreaks. It explored that the number of the incidence of Wuhan City had a rapid upward trend. In the early stage, except for a sudden sharp increase on Jan 27, daily incidence had been increasing as of Feb 4. And from this day onwards, there was a flat phase of volatility. The downward trend began on Feb 11. The other two regions, Hubei Province exclude Wuhan and China exclude Hubei Province were showing roughly similar trends. They first slowly increased to a certain level and then decreased slowly, and the peaks were significantly lower than those in Wuhan.

**Figure 3.**
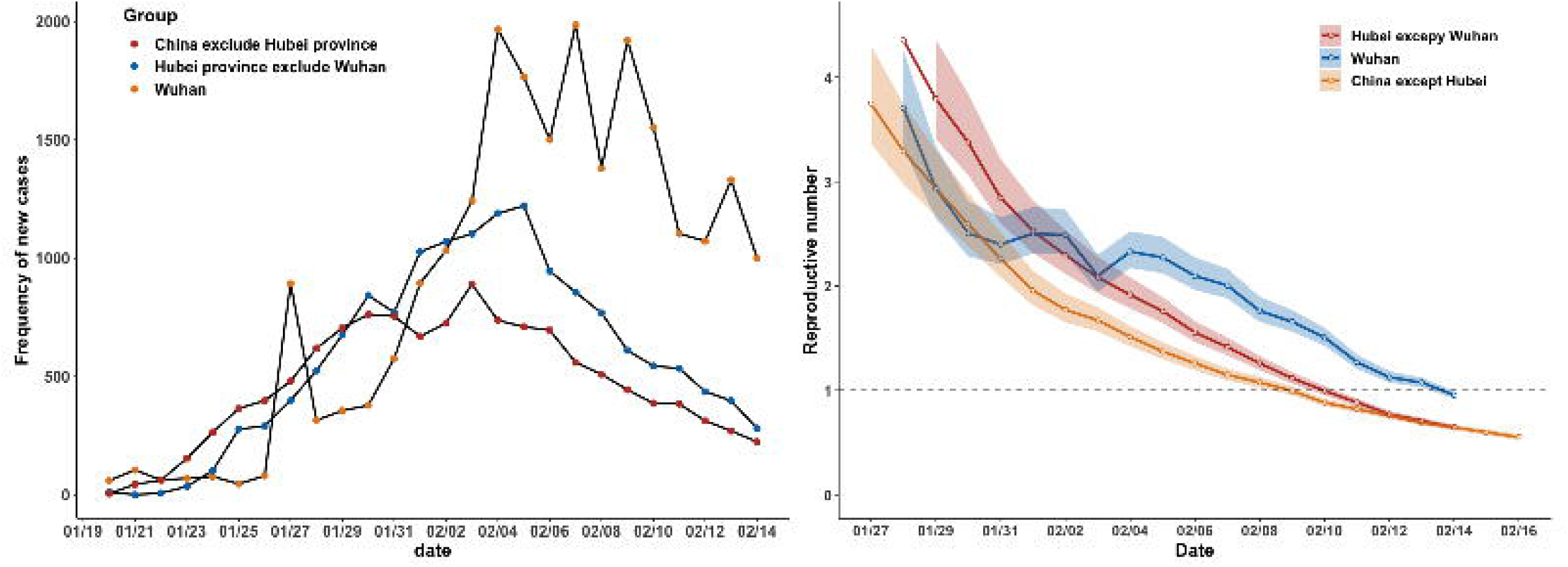
The daily incidence and R_t_ in three areas with varying levels of China. Figure 3A showed the daily incidence in three areas with varying levels; Figure 3B showed the R_t_ in three areas with varying levels.

Figure 3B revealed the Rt of Wuhan, where the epidemic of COVID-19 originate as well as the most sever epidemic, showed a dynamic change and a downward trend. And the R_t_ (1.06±0.02) was close to one on February 13th, and dropped below one (0.96±0.02) the next day. China excluded Hubei Province took the lead in dropping below one (0.99±0.02) on February 9, followed by Hubei Province excluded Wuhan the next day (0.99±0.02). In addition, we compared the R_t_ among each province in China, the results were shown in the section two of Append S1. The more detail of R_t_ result were shown in third sheet of S2 Table.

### Estimating the foreign countries’ R_t_

The outbreak had spread many countries around the world, of which Japan, Singapore, Korea, Italy and Iran were the most serious. Therefore, we also analyzed the changed of the R_t_ in these countries as well as other overseas countries and regions to find out the international spread of COVID-19. During February 7 (0.92±0.26) and February 13(0.46±0.23), the R_t_ in Japan were below one, which meant the COVID-19 was not spread widely. However, since February 14 the Japan’s R_t_ showed a suddenly growth trend and even increased to 3.26±0.63 on February 16. And Korea also showed the similar change of R_t_ with Japan, but greater than Japan’s R_t_. The epidemic in Singapore was slightly better. Due to less reported cases data, the estimating for R_t_ had a wide range, but got stable estimates in the later stages. Compared with Japan, Singapore’s R_t_ fluctuated below two, and the current R_t_ showed that it had fallen to one on February 21 (0.92±0.17), indicating the epidemic was under control. Another two countries (Iran and Italy) also had a fluctuating R_t_, which was always greater than one(4.45±0.57 and 5.40±0.63 as February 26). But other overseas countries showed the R_t_. had fallen to one recently (0.89±0.06 on February 23). (Figure 4) The more detail of result was shown in forth sheet of S2 Table.

**Figure 4.**
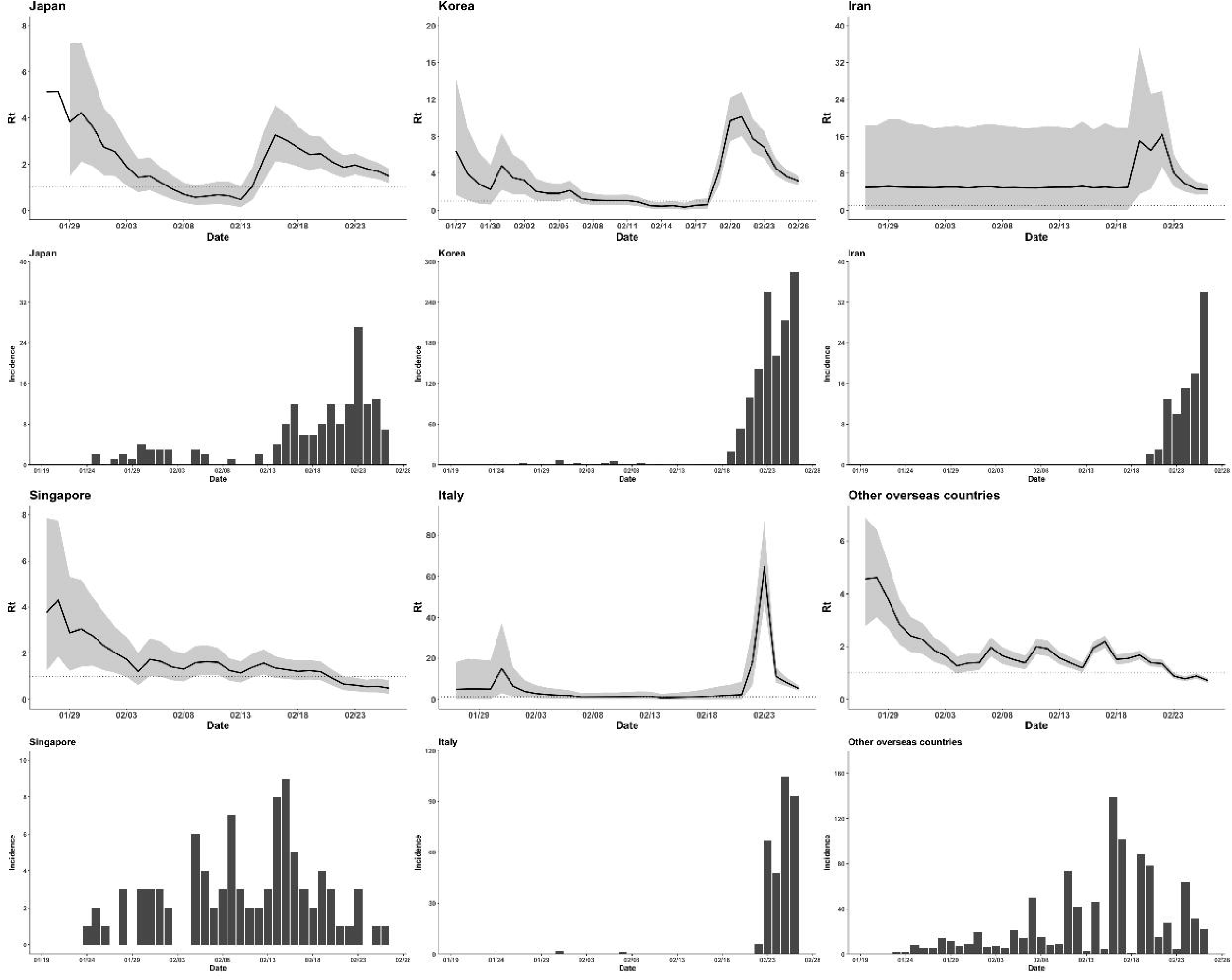
The distribution of daily incidence and R_t_ in top five sever outbreak countries (excluding China) and combined other overseas countries. The histogram plots showed the daily incidence of each country; the line plots showed the R_t_ of each country.

## Discussion

In this study, we mainly applied Thompsona’s method to estimate the R_t_ of COVID-19 in different areas to assess the spread of the epidemic in China and oversea. We found out that the R_t_ in China had almost been falling below one, which mean the non-pharmaceutical intervention in China were effective. But the situation of overseas is not optimistic as R_t_ >1, which indicated the epidemic was still in the stage of gradual spread.

As of Feb 29, almost 80,000 confirmed cases of COVID-19 in China have been reported and nearly 3,000 people have died. As we all know, Wuhan as the origin outbreak is also the most severely affected area accounting nearly 60 percent of cumulative cases nationwide in China. And the fatality rate in Wuhan is much higher than in other regions.^14^ Although existing confirmed cases was still increasing the rate of increase was steadily declining. It meant the series of non-pharmaceutical interventions implemented by the Chinese Government have achieved remarkable achievement. In this study, we found daily incidence of three areas were at different levels in China. Most of the new cases still came from Wuhan. The overall daily incidence has been gradually decreasing since February 4 and the daily incidence in Hubei Province excluded Wuhan and China excluded Hubei Province was gradually approaching zero, while Wuhan was still at a relatively high level and the decline of incidence was relatively slow.

As of Feb 11, the cumulative number of reported cases in China showed a bifurcation. According to the update of “The diagnosis and treatment program of novel coronavirus pneumonia (Trial Fifth Edition)”,^15^ only Hubei province added the classification of “clinical diagnosis”, that the suspected cases with clinical features of pneumonia were clinically diagnosed cases. And it was cancelled in the sixth edition a few days. That’s because the Chinese authorities considered the limited medical resources and huge number of suspected patients. During Feb 4^th^ and Feb 10^th^, the daily incidences in Wuhan fluctuated around 2,000, indicating that due to limited medical resources, many suspicious patients were still waiting for diagnosis, which might result in aggravated conditions and the risk of potential outward transmission. To speed up controlling the epidemic, Chinese authorities had adopted the standard of adding clinical diagnosis. When most suspected patients are diagnosed and treated effectively, the chain of disease transmission will become clearer.

In order to find out the trend of the epidemic, we calculated the R_t_ in different regions in China. And we also further calculated the R_t_ abroad to understand the international development of the epidemic. Based on official data and published literature, we analyzed the interval time between the onset dates of 126 pairs of infected and infected cases. By fitting the Gamma distribution, we got the median serial time of COVID-19 is 4.18 days. It’s longer than Hiroshi Nishiura’s result (2.6 days), which estimated by 26 infector-infected pairs.^16^ The intergenerational time obtained in this study was smaller than the estimated value of 5 clustered outbreaks in 425 cases at beginning of the epidemic. And the time was also smaller than the average incubation period of 5.2 days, which was also estimated by 425 cases at beginning.^12^ We supposed that the infected person was infected before the onset of symptoms.

Then, by inputting the serial time distribution and the latest (as of February 16) the incidence of laboratory confirmed cases with 7 days as the moving time window to estimate the value of R_t_.

At first, we compared the R_t_ among three different regions in China. We could see all R_t_ in three areas had downward trends, as well as had fallen below one. It meant the series non-pharmaceutical interventions implemented in China had achieved some achievement.

And Wuhan as the origin of the epidemic, which also having the majority of patients, had the different change trend of R_t_. Compared with other two regions, the average R_t_ of Wuhan was higher and token longer time to drop below one. In addition, as we all know, there are still over thousands accumulated cases in Wuhan, so it still has a long way to control the epidemic.

Optimistically, the contagiousness of COVID-19 in China has been well controlled. As long as strict management is maintained, the number of incidences will drop to zero soon. As of February 25, the incidence in China had dropped from thousands to hundreds, meanwhile the number of cures was also increasing.^17^ Zeliang Chen et al predicted the peak incidence to be in early or middle February.^18^

Secondly, we assessed the development of the epidemic overseas. Considering the extent of the epidemic, we listed top five countries with relatively severe outbreaks, Japan, Korea, Singapore, Iran and Italy, and combined other overseas countries for R_t_ calculations. As of February 26, except Singapore, other four countries’ R_t_ were still upper than one, which meant the epidemic might be growing abroad. On February 7, the Singapore government officially decided to upgrade the response level of the outbreak response system from yellow to orange in view of the discovery of several local cases in Singapore, which were unrelated to previous cases and had no history of travel to China.^19^ In our study, the result showed the R_t_ in Singapore had fallen below to one on February 20, which meant the outbreak in Singapore had been under control. Although the other overseas countries’ R_t_ also had fallen below to one, it didn’t mean the outbreak around the world was in control. On 30 January 2020, the outbreak was declared a Public Health Emergency of International Concern by WHO. And WHO and countries have been engaged in massive preparedness activities.^20^ And according to the growth rate of the worldwide new confirmed cases, there were still a trend of rapid growth, which liked the early stage of epidemic in Wuhan. We hope it should be taken more attention and learn the experience from China or Singapore.

## Limitations

Our study used the latest generation time and new confirmed case data at the same time, trying to estimate the R_t_ of each region in China and oversea, which could provide a basis for grasping the development trend of the epidemic and evaluate prevention and the interventions. But there were some limitations in our study. First, we collected the personal history of epidemiological investigation data from different publish websites to estimate the serial interval, which made bias. We hope the government might build the database to collect the history epidemiology data, what could provide more reliable information.^21,22^ Second, failure to exclude imported cases in the calculation of R_t_ might make bias, but it could still reflect a certain epidemic situation and the effectiveness of prevention and intervention. Another limitation is that we used the time-series data of confirmed cases to calculate according to the official date of diagnosis. So, the date we estimated that the R_t_ went below one should be delayed by about 10~17 days (the incubation period of 7 days added days from onset to diagnosis of 10 days). Although we estimated the time when the epidemic was under control had a delay than the actual time, it could still provide information of the trend of the epidemic.

## Conclusions

In conclusion, the outbreak in China has been well controlled, but the worldwide pandemic has not been well controlled. However, there is still uncertainty about what the SARS-CoV-2 virus will do worldwide and even in China after the country inevitably lifts some of its strictest control measures.

## Data Availability

The data access committee comprises four corresponding authors and there is no restriction to data access.

https://www.who.int/emergencies/diseases/novel-coronavirus-2019/situation-reports

## Acknowledgments

L Zheng and K Qin contributed equally to this manuscript. L Zheng, K Qin conceived and designed the study. L Zheng wrote the paper. X Chen, S Huang and D Liu collected data. W Liao reviewed and edited the manuscript. H Liang guided the study. All authors read and approved the manuscript. We thank the data sharing from the China Centre for Disease Control and Prevention.

## Supporting information

**S1 File. The method to estimate R_t_ and the result of comparing all provinces’ changing of R_t_ in China**.

The word file named “S1 File” included two section: A. The method to estimate the time-dependent reproduction number (R_t_); B. The result of comparing all provinces’ changing of R_t_ in China.

**S1 Table. The detail information of 126 infector-infected pairs**.

**S2 Table. The detail data of the result in China**.

The excel file named “S1 Table” included four sheets. The data were mainly corrected from Chinses CDC’s official data were shown in the first sheet named “China”; the incidence in three region in China were shown in the second sheet named “incident”; and the result of R_t_ in China and outside China respectively in the third and fourth sheet.

